# Validation of Left Ventricular High Frame Rate Echo-Particle Image Velocimetry against 4D Flow MRI in Patients

**DOI:** 10.1101/2023.11.27.23298719

**Authors:** Yichuang Han, Daniel J. Bowen, Bernardo Loff Barreto, Robert. R. Zwaan, Mihai Strachinaru, Rob J. van der Geest, Alexander Hirsch, Annemien E. van den Bosch, Johan G. Bosch, Jason Voorneveld

## Abstract

**Aims:** Accurately measuring intracardiac flow patterns could provide insights into cardiac disease pathophysiology, potentially enhancing diagnostic and prognostic capabilities. This study aims to validate Echo-Particle Image Velocimetry (echoPIV) for *in-vivo* left ventricular intracardiac flow imaging against 4D flow MRI.

**Methods and Results:** We acquired HFR contrast-enhanced ultrasound images from three standard apical views of 26 patients who required cardiac MRI. 4D flow MRI was obtained for each patient. Only echo image planes with sufficient quality and alignment with MRI were included for validation. Regional velocity, kinetic energy and viscous energy loss were compared between modalities using normalized mean absolute error, cosine similarity and Bland-Altman analysis.

Among 24 included apical view acquisitions, we observed good correspondence between echoPIV and MRI regarding spatial flow patterns and vortex traces. The velocity profile at base-level (mitral valve) cross-section had cosine similarity of 0.92 ± 0.06 and normalized mean absolute error of 14 ± 5%. Peak spatial mean velocity differed by 3 ± 6 cm/s in systole and 6 ± 10 cm/s in diastole. The kinetic energy and rate of energy loss also revealed a high level of cosine similarity (0.89 ± 0.09 and 0.91 ± 0.06) with normalized mean absolute error of 23 ± 7% and 52 ± 16%.

**Conclusions:** Given good B-mode image quality, echoPIV provides a reliable estimation of left ventricular flow, exhibiting spatial-temporal velocity distributions comparable to 4D flow MRI. Both modalities present respective strengths and limitations: echoPIV captured inter-beat variability and had higher temporal resolution, while MRI was more robust to patient BMI and anatomy.

## Introduction

Quantification of intracardiac flow patterns may support diagnosis and understanding of various cardiac conditions, and assessment of underlying pathophysiology. Such information may help risk assessment, including predicting thrombus formation and gauging treatment effectiveness, as well as informing potential future treatment options. Recent studies suggested that cardiac hemodynamics are correlated to different types of cardiomyopathy [1, 2, 3, 4], including dilated cardiomyopathy[5][6]. At present, 4D flow MRI is the gold standard of intracardiac flow imaging, offering detailed 3D velocity vectors throughout the cardiac cycle for comprehensive quantification and visualization of cardiac blood flow patterns. However, it has practical limitations such as high costs, long scan durations, and incompatibility with certain patient conditions or implants.

Echocardiography is a readily accessible, real-time imaging technique used extensively in clinical practice. Doppler ultrasound techniques enable the measurement of blood flow velocity through the valves[7][8], but only provides velocity information along the ultrasound beam direction, limiting assessment of complex, multi-directional flow patterns.

Echo-Particle Image Velocimetry (echoPIV) provides a 2D velocity field, revealing intracardiac hemodynamics[9][10]. EchoPIV based on conventional (low frame rate) imaging has explored the relationship between flow and heart diseases but is limited in quantifying the full velocity spectrum due to frame-rate limitations[11][12][13]. High frame rate (HFR) echoPIV employs HFR contrast-enhanced ultrasound imaging to estimate blood flow patterns. HFR imaging uses diverging wave transmissions and software beamforming and allows imaging at 100× the frame rate of conventional echocardiography[14].

In previous studies, echoPIV has been compared with optical PIV (*in-vitro*)[15] and pulse-wave Doppler (*in-vivo*)[6][16][17]. However, an *in-vivo* validation for the whole intracardiac velocity field was still absent. Therefore, this study primarily aims to validate 2D echoPIV against 4D flow MRI for *in-vivo* left ventricular flow imaging. Additionally, we examine the advantages and disadvantages of both imaging modalities. Finally, we report disturbed flow patterns in a patient with a severely dilated left ventricle (LV) and another one with left bundle branch block.

## Methods

### Setup of Study and Patient Selection

Patients referred for cardiac MRI for screening of cardiomyopathy or determining the underlying etiology were selected. Exclusion criteria were hypertrophic cardiomyopathy, non-sinus rhythm, and poor acoustic windows in conventional echocardiography. HFR echoPIV was acquired on the same day as cardiac MRI (except for one patient). This study was approved by the Erasmus MC Medical Ethic Review Committee (Rotterdam, the Netherlands), and written informed consent was obtained from all patients (METC-2018-057, NL63755.078.18).

### 4D Flow MRI

Image acquisition was performed on a 1.5 T clinical MRI scanner (SIGNA Artist, GE Healthcare, Milwaukee, WI, USA) using an anterior phased-array coil. The protocol included breath-hold balanced steady state free precession (bSSFP) cine images in the three standard long-axis views and a contiguous stack of short-axis images, all with 30 phases per cardiac cycle. Furthermore, a free-breathing, retrospectively ECG-gated 4D flow sequence with parameters shown in Table 1 was acquired directly after administration of gadolinium contrast agent (Gadovist 0.2 mmol/kg) prescribed in the axial plane covering the whole heart.

**Table 1.** Technical parameters of 4D flow sequence.

|  |  |
| --- | --- |
| Acceleration method | HyperKat factor 6 with compressed sensing |
| Flip angle (degrees) | 15 |
| Arrhythmia rejection | 30% |
| Respiratory compensation | 20% |
| Field of view (mm) | 380 |
| Phase field of view | 60-100% |
| Repetition time (ms) | 4.2 |
| Number of excitations | 4 |
| Velocity encoding (cm/s) | 150 |
| Reconstructed number of phases | 30 |
| Acquired resolution (mm<sup>3</sup>) | $2.4 \times 2.4 \times 2.4$ |

The 4D flow processing was performed using MASS software (Leiden University Medical Center, Leiden, the Netherlands)[18]. A second-order plane fit was applied as a static-tissue interpolation offset correction to correct for phase-offset errors. The contours of the LV were delineated on 2D anatomical MRI sequences. To ensure precise alignment, the anatomical positions of these 2D slices were extracted from the DICOM data and were used to define matching cut-planes in the 4D Flow MRI volume. Flow data corresponding to these planes were subsequently exported from the 4D sequence and further processed using Matlab (R2019a, MathWorks, Natick, MA, USA).

### Echocardiography

The echocardiographic protocol consisted of two parts: a standard clinical protocol using a clinical ultrasound machine (EPIQ 7 with probe X5-1, Philips Healthcare, Best, The Netherlands); and the HFR contrast-enhanced ultrasound recordings using a research ultrasound machine (Vantage256, Verasonics, Kirkland, WA, USA) with a phased-array probe (P4-1, ATL, Bothell, WA, USA). The setup overview is shown in Figure 1. The clinical protocol includes B-mode and colour Doppler acquisitions in the apical two, three and four-chamber (A2C, A3C & A4C) views and pulsed-wave Doppler measurements at the mitral valve tips and the LV outflow tract (LVOT).

**Figure 1.**
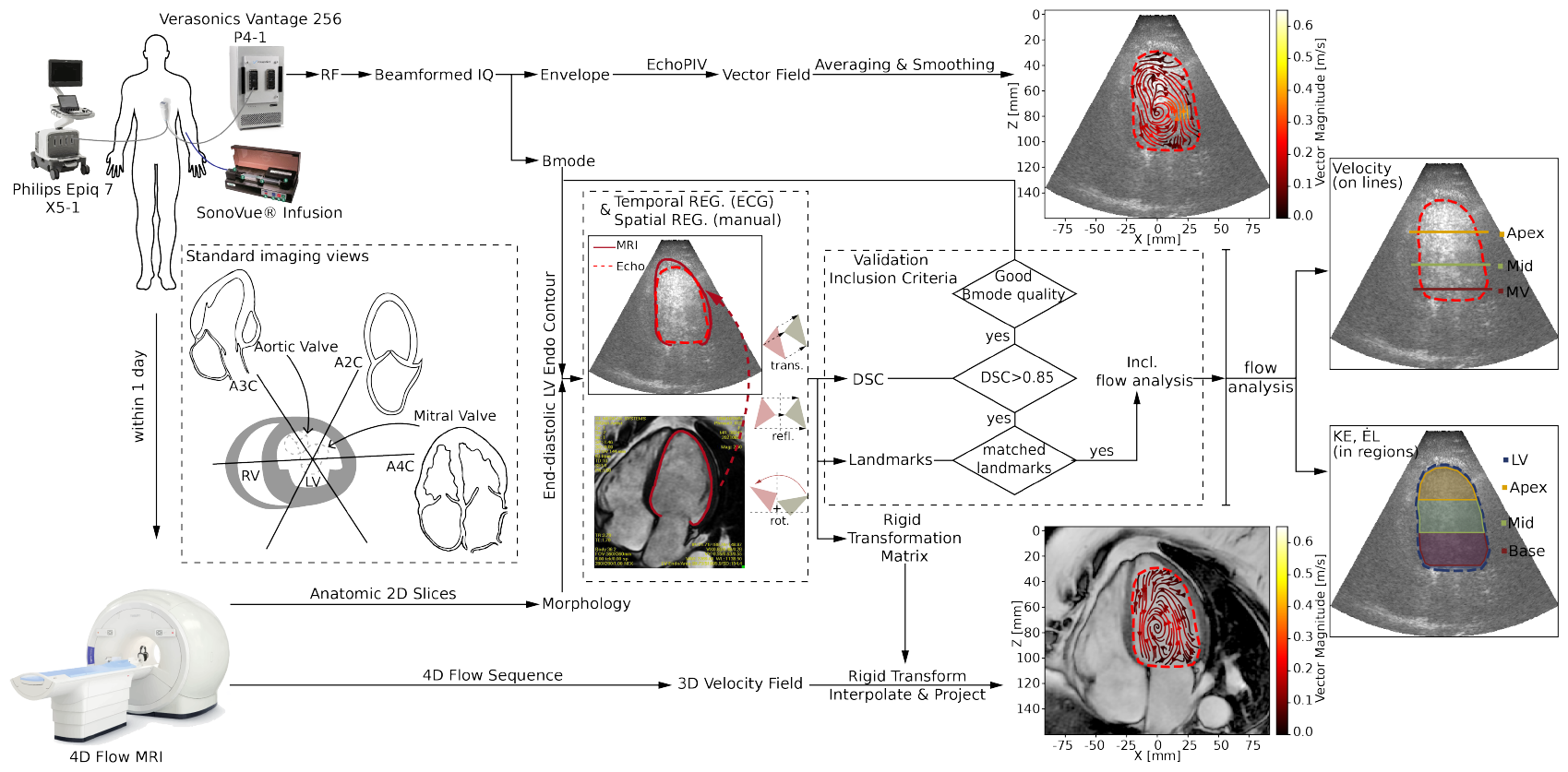
Overview of the experimental setup, signal processing and flow analysis pipeline. DSC: Dice coefficient; KE: Kinetic energy; E·L: Rate of energy loss; LV: Left ventricle; REG: registration

Following the clinical imaging protocol, a diluted solution (1:3) of ultrasound contrast agent (SonoVue, Bracco Imaging SpA, Milan, Italy) in saline was intravenously infused (1ml/min) using a continuous infusion pump (VueJect BR-INF 100, Bracco Imaging SpA). Upon verification of contrast agent arrival in the LV, the imaging system was switched to the research ultrasound machine for HFR imaging. HFR data were obtained and beamformed with the setup described by Voorneveld et al.[16], a detailed description can be found in supplementary data. The pulse repetition frequency was chosen to reach the physical limitation of imaging depth (3553 − 6012Hz). Each acquisition captured a total of 3000 frames of data (∼ 2.5 seconds), allowing for the recording of at least two cardiac cycles.

### EchoPIV Processing

EchoPIV was performed using custom Matlab code (R2022a, MathWorks, Natick, MA, USA) and, in brief, involved subdividing each image into smaller blocks and tracking the HFR contrast-enhanced ultrasound speckle pattern over time to obtain the velocity information in each block. A more technical description is provided in the supplementary data[15][16]. The final frame rate (after pulse inversion, angular compounding and ensemble averaging) of the velocity data was ∼ 122.5 frames/second. The vector field obtained by echoPIV was interpolated onto a uniform cartesian grid for flow analysis.

### Registration

For an accurate comparison of MRI and ultrasound flow fields, spatial and temporal registration between the modalities is required. Temporal registration involved up-sampling the time series of MRI measurements (30 frames/cardiac cycle) to match the frame rate of the ultrasound images (∼100 frames/cardiac cycle). The R peaks of the ECG signals were used to separate echocardiographic acquisitions into cycles and to synchronize them with the retrospectively ECG-gated MRI data.

Spatial registration: MRI LV endocardial contours were manually drawn on the standard bSSFP long-axis cine images in each phase of the cardiac cycle using MASS. These contours were then spatially aligned (manually) to the ultrasound B-mode image by using a rigid transformation. The same transformation was applied to the MRI vector field and the transformed 3D vectors were interpolated on the same grid as the ultrasound data and projected onto the 2D ultrasound plane.

### Feasibility and Assessment of Registration

Inclusion criteria consisted of two aspects: 1) B-mode image quality, and 2) co-registration of MRI and echocardiography. B-mode image quality indicates the clinical feasibility of echoPIV for flow estimation. Acquisitions with poor contrast/signal-to-noise ratio, blurred point spread function, or bubble signal loss in the basal region were considered unfeasible and excluded from flow comparison.

Goodness of registration was quantitatively evaluated using the Dice coefficient between the end-diastolic LV endocardial MRI and ultrasound contours. We first excluded the acquisitions with severe foreshortening by setting a threshold on the Dice coefficient(> 0.85). Subsequently, we performed a visual assessment of all views by multiple independent observers (YH, JV, JGB) followed by a consensus session selection. This evaluation involved a careful visual comparison between the two modalities of key anatomical landmarks (the apical shape, septal wall, mitral valve and aortic valve, LVOT, and the position of papillary muscles) and the possible plane mismatch. The flowchart of the validation inclusion process is shown as part of Figure 1.

### Comparison of Flow between EchoPIV and 4D Flow MRI

We evaluated the spatial-temporal agreement of the two techniques using their velocity profiles, and commonly derived regional flow parameters: kinetic energy (KE), rate of energy loss (E·L) and circulation, which are defined in the supplementary data.

The LV was split into basal, mid, and apical regions by first selecting two basal points on the mitral anulus and a third point on the LV apex. The apex and the mid-point of the two basal points define the LV long axis. Two lines parallel to the line connecting the two basal points were then constructed that trisected the LV long axis, effectively dividing the LV into three segments: the base, mid, and apex regions (see Figure 1). In addition to these, we identified another parallel line at 1/9 of the LV long axis length for the mitral valve inflow. The velocity profiles were evaluated on the three parallel lines mentioned above - inflow, mid, and apex. The flow comparison was performed over the whole LV as well as locally (in the basal, mid, and apical regions of the LV). Two quantitative measures were used for comparison: Cosine Similarity (to assess similarity in the shape of the temporal pattern of each parameter between modalities) and Normalized Mean Absolute Error (NMAE, to assess absolute differences between parameters measured by each modality).

### Statistical Analysis

Statistical analysis was performed using Matlab Statistics and Machine Learning Toolbox. To evaluate the clinical characteristics of all study participants against those of the comparison group, a Student’s t-test was conducted for the continuous data and a Fisher’s exact test was conducted for the categorical data. P-values < 0.05 implied statistical significance. Student’s t-test was also applied to compare differences in Dice coefficient across three apical views. Bias and limits of agreement for temporal peak velocities between modalities, during systole and diastole, were estimated using Bland-Altman analysis[20], reported with 95% confidence intervals (95% CI).

## Results

### Feasibility and assessment of registration

As shown in Figure 2, among 78 standard apical view HFR acquisitions from 26 patients, 61 (78%) had sufficient B-mode image quality for echoPIV flow estimation and thus could be considered clinically feasible for echoPIV processing.

**Figure 2.**
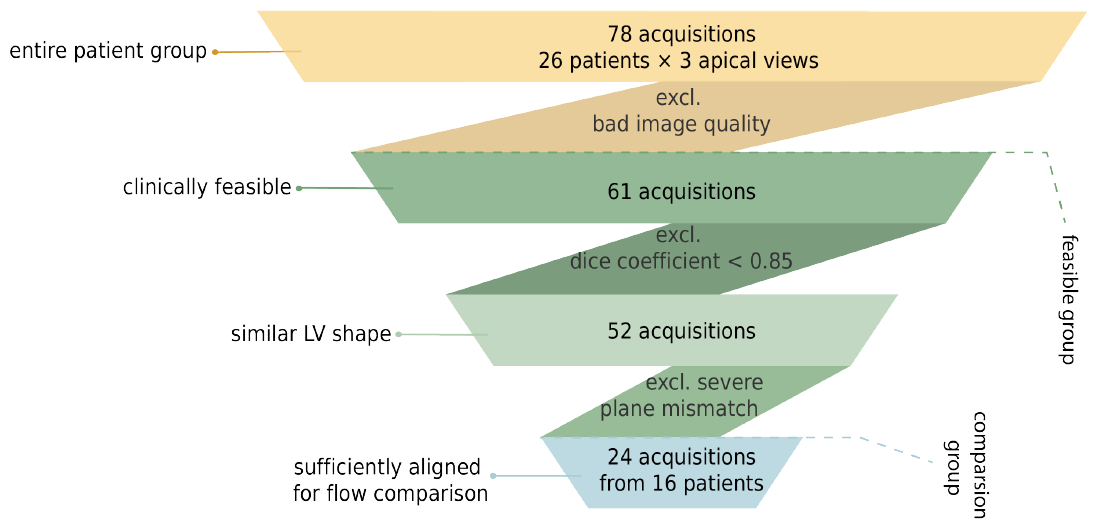
Flow chart for patient inclusion for quantitative flow comparison

The mean Dice coefficient for all 78 acquisitions was 0.88 ± 0.05, indicating a high-level similarity between the end-diastolic echocardiographic and MRI masks. Dice coefficients for A4C, A3C and A2C (v.s. all views) showed no significant differences among the three types of standard apical views.

After removal of acquisitions that did not meet the stringent alignment criteria (Movie S.6, S.7 and S.8 show examples of removed acquisitions), 24 acquisitions from 16 patients were selected for the quantitative flow comparison: 12 A4C, 6 A3C, and 6 A2C views.

### Hemodynamic and Echocardiographic Data

General clinical, conventional echocardiographic and MRI characteristics for all patients and those included for comparison are displayed in Table 2. No significant differences were found between the comparison group and the total patient group.

**Table 2.** Clinical Information of the patients.

| Parameters | All patients (n = 26) | Comparison group (n = 16) | p-value |
| --- | --- | --- | --- |
| ***General characteristics*** |  |  |  |
| Age | 46 [19-76] | 39 [21-77] | 0.28 |
| Sex (Male/Female) | 17/9 | 9/7 | 0.74 |
| BMI | 24.8 (20.7-27.1) | 24.8 (20.2-26.7) | 0.69 |
| Heart rate(bpm) | 66 (57-74) | 72 (65-78) | 0.28 |
| Systolic blood pressure(mmHg) | 123 (118-135) | 125 (118-136) | 0.63 |
| Diastolic blood pressure(mmHg) | 73 (67-79) | 74 (67-77) | 0.84 |
| ***Echocardiographic characteristics*** |  |  |  |
| 2D LV end-diastolic volume (ml) | 187 (148-230) | 180 (133-198) | 0.93 |
| 2D LV end-systolic volume (ml) | 96 (72-128) | 81 (58-96) | 0.88 |
| LV end-diastolic diameter (mm) | 58 (54-62) | 56 (48-62) | 0.65 |
| Peak velocity E (m/s) | 0.7 (0.6-0.8) | 0.8 (0.6-0.9) | 0.19 |
| Peak velocity A (m/s) | 0.6 (0.5-0.7) | 0.6 (0.5-0.7) | 0.57 |
| E/A | 1.0 (0.8-1.4) | 1.3 (1.0-1.5) | 0.25 |
| E/e' | 7.5 (6.4-10.0) | 7.1 (6.4-11.5) | 0.92 |
| Peak velocity LVOT (m/s) | 1.0 (0.6-1.3) | 1.0 (0.6-1.3) | 0.84 |
| LV ejection fraction (%) | 53 (40-56) | 55 (49-59) | 0.33 |
| ***MRI characteristics*** |  |  |  |
| LV end-diastolic volume (ml) | 204 (166-239) | 195 (164-226) | 0.92 |
| LV end-systolic volume (ml) | 120 (72-128) | 81 (68-122) | 0.92 |
| LV ejection fraction (%) | 52 (41-55) | 52 (46-58) | 0.47 |
| LV ejection < 45% (n) | 10 | 4 | 0.51 |
| LV Mass (gr) | 126 (104-155) | 121 (93-137) | 0.45 |

Data expressed as median (interquartile range) or number; age expressed as average [range].
LV=left ventricular; LVOT=left ventricular outflow tract; BMI=body mass index; E=early mitral inflow velocity; A=late mitral inflow velocity; e' = mitral annular early diastolic velocity

### Flow Comparison: An Exemplary Case

Figure 3 shows the flow estimation results of echoPIV and 4D flow MRI for one exemplary patient. Figure 3.A and Figure 3.B show the flow streamlines during the formation of the vortex ring at ejection and early filling respectively (corresponding animation in Movie S.1).

**Figure 3.**
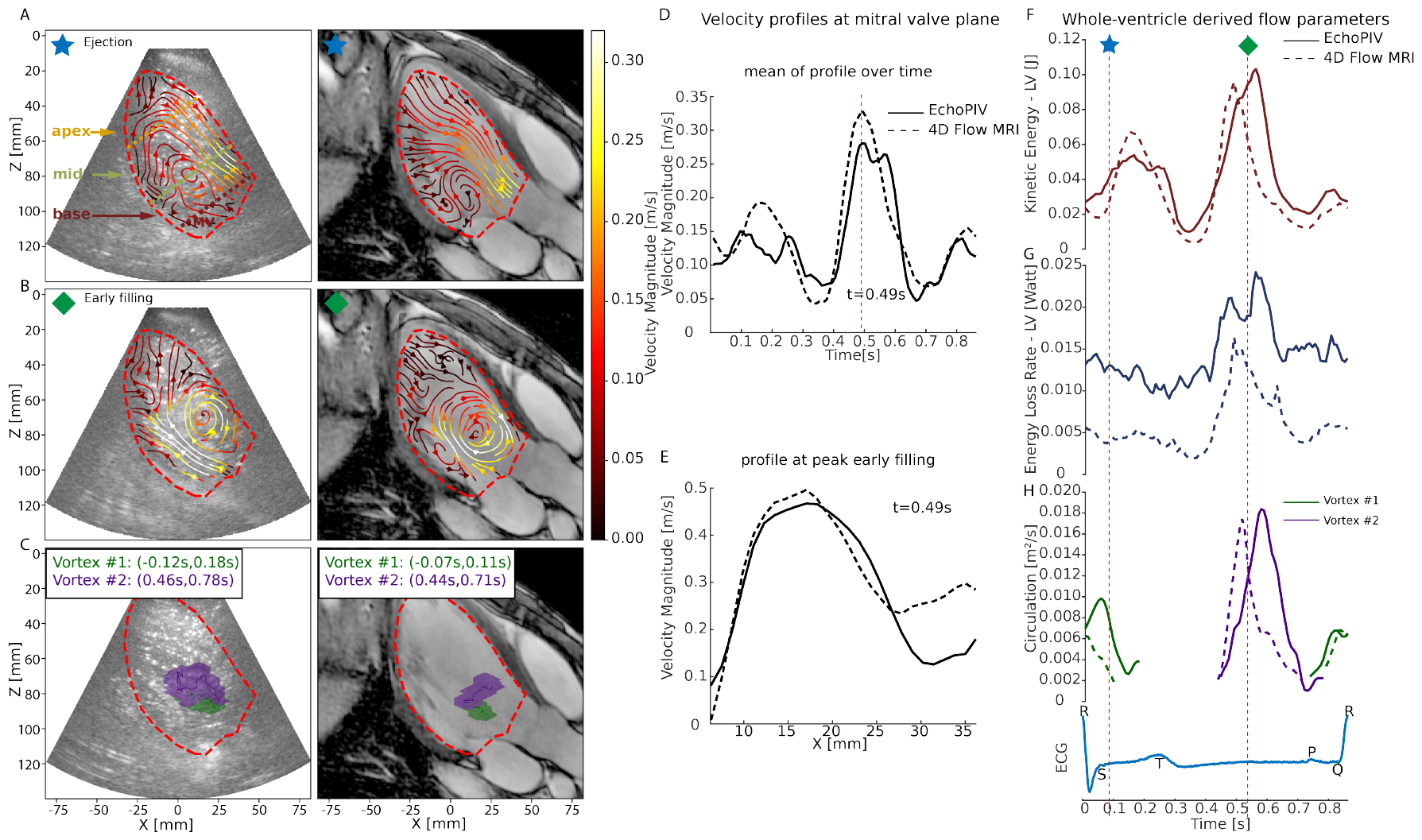
(A) Flow streamlines of EchoPIV (left) and MRI (right) at *t* = 88*ms* (ejection phase). (B) Flow streamlines of EchoPIV (left) and MRI (right) at *t* = 537*ms* (early filling). (C) Vortex traces of EchoPIV (left) and MRI (right). Velocity profiles at mitral valve plane: (D) mean of profile over time, (E) profile at peak of early filling. Whole-ventricle derived flow parameters: (F) kinetic energy, (G) rate of energy loss, (H) circulation, ECG as time reference with star and diamonds marking the ejection and early filling

The comparison of the main vortex traces is shown in Figure 3.C (Movie S.2). Figure 3.D and E show the spatial mean velocity and the spatial velocity profile at peak inflow along the inflow line (the red dashed line at the mitral valve in Figure 3.A). The parameters derived from the velocity field (KE, E·L and circulation in the detected vortex region) are shown in Figure 3.F, with ECG R peak as a time reference.

### Agreement between Echocardiographic- and MRI-Flow Parameters

The velocity profile analysis for the comparison group (Figure 4.A and C), indicated a good agreement across all three levels.

**Figure 4.**
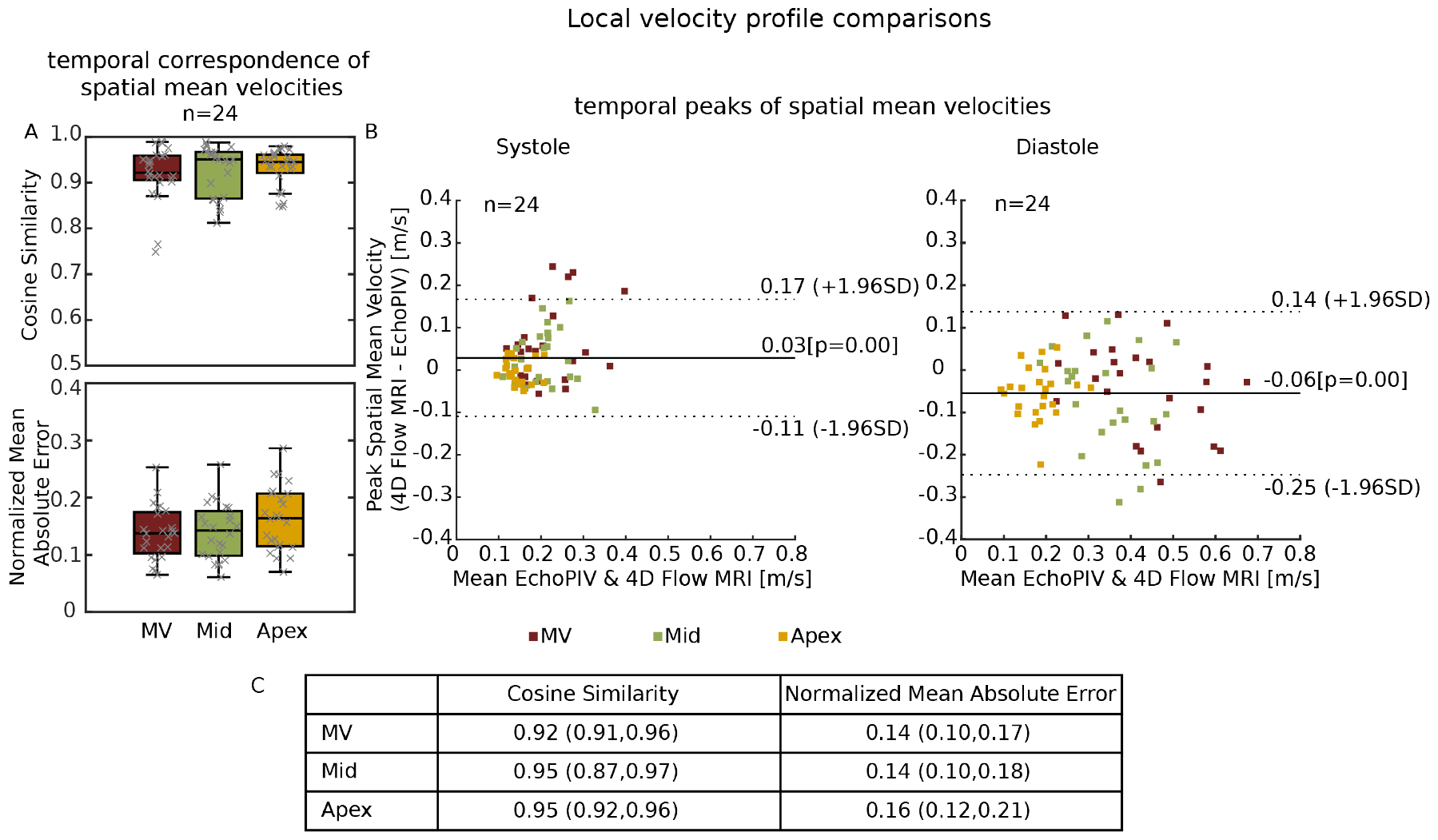
(A) Box plot of cosine similarity and normalized mean absolute error of the spatial mean velocity profile through lines at mitral valve (MV) inflow, mid and apex level. Black lines in the middle indicate median, boxes extend to 25th and 75th percentiles, and whiskers indicate the range excluding samples outside of 1.5 times the interquartile range. (B) Bland-Altman plot of the peak spatial mean velocity through lines at MV inflow, mid and apex level (as demonstrated in Fig 1.A) during systole and diastole. (C) Table summarizing the box plot in Fig. 4.A, data as median(25th, 75th percentiles)

Bland-Altman analysis of peak spatial-mean velocity (Figure 4.B) revealed that echoPIV measured higher velocities than MRI during diastole and lower velocities during systole. Moreover, echoPIV exhibited higher velocities in apex than MRI.

As shown in Figure 5, the median cosine similarity for regional KE and E·L lay in the range of (0.82,0.91) and (0.88,0.92). The NMAE exhibited higher values than those of velocity analysis, with its median lying in the range of (0.22,0.36) and (0.45,0.53) for KE and E·L. The apical region exhibited a larger difference between echo and MRI than the whole LV, characterized by a lower cosine similarity and higher NMAE.

**Figure 5.**
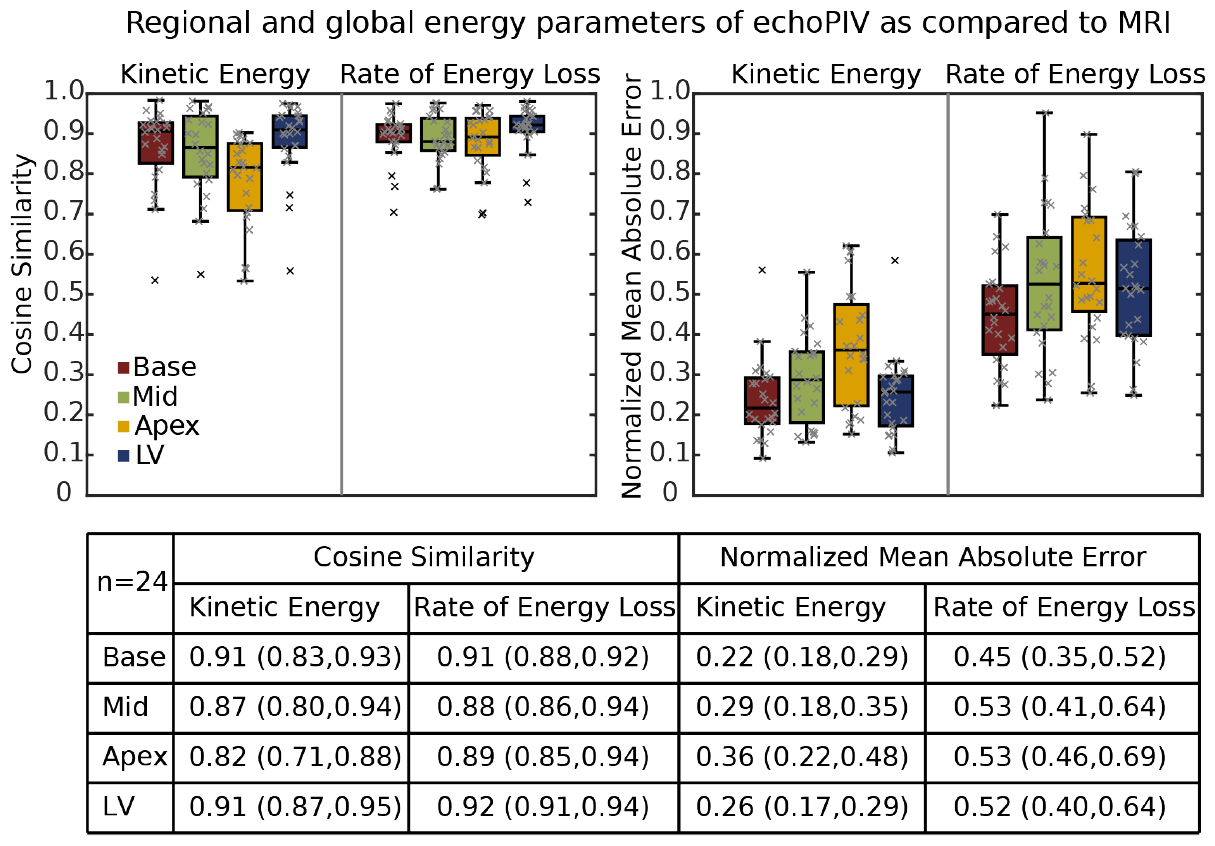
Box plot of cosine similarity and normalized mean absolute error of kinetic energy and energy loss rate in base, mid, apex regions and the whole left ventricle (LV). Data as median(25th, 75th percentiles)

When comparing the results of all 61 clinically feasible acquisitions to the inclusion group, a higher variance in the Bland-Altman analysis for velocity profiles (Figure S.1) and a slightly declined level of agreement for KE and E·L (Figure S.2) was observed.

### Inter-beat Variability

Ultrasound allowed an inter-beat variability analysis. Although only patients with sinus rhythm were included to ensure high-quality MRI results, we still observed considerable inter-beat variability in the hemodynamics with echoPIV between cardiac cycles for certain patients.

A representative example is provided in Figure 6. Variability in flow streamlines (Figure 6.A, Movie S.3) and energy parameters (Figure 6.B) were observed over three consecutive cardiac cycles.

**Figure 6.**
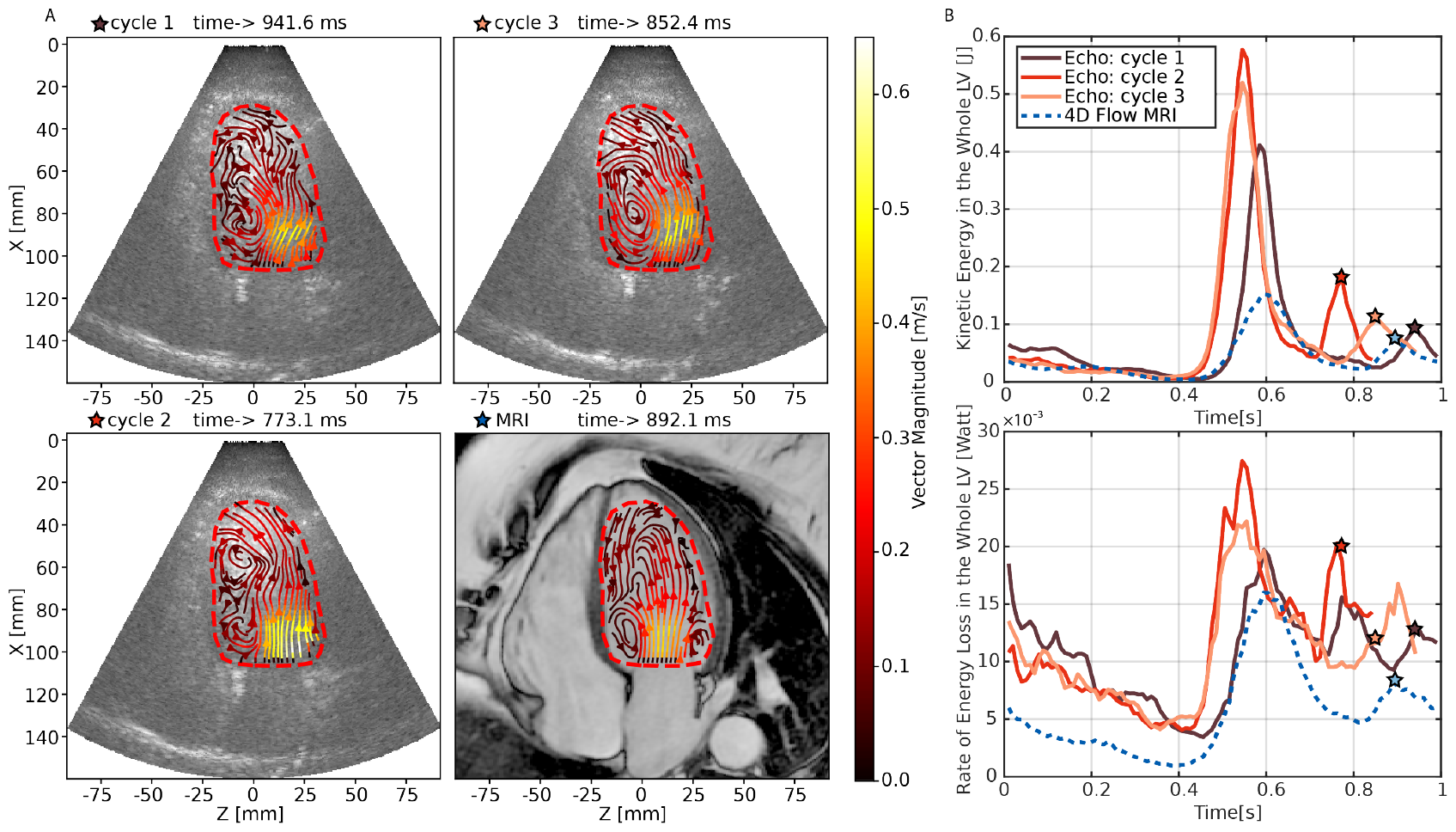
Example of inter-beat variability captured by echoPIV: (A) Streamline of the blood flow at peak late filling of each cycle of echoPIV and MRI measurement. (B) Kinetic energy and rate of energy loss in the whole left ventricle of each cycle of echoPIV and MRI measurement

## Discussion

For the patients with sufficient image quality (78%), using the echoPIV flow estimation, we can obtain 2D vector velocity field information equivalent to 4D flow MRI. Combined with echo’s low cost plus its ability to assess inter-beat variability, HFR echoPIV may be useful for follow-up examinations of patients and guiding treatment strategies.

### Qualitative Comparison

Qualitatively, similar flow patterns were observed between echoPIV and MRI, especially during diastole where there was striking resemblance between the filling flow patterns and velocity magnitudes. In systole there was a weaker agreement, where the outflow jet velocities were underestimated in many echoPIV acquisitions (Figure 4.B). This underestimation has also been observed in previous studies[16][21] and may be due to several reasons: 1) slight deviations of echo imaging plane from the LVOT plane, 2) signal decorrelation occurring during the pulse inversion process, 3) strong spatial velocity gradients in the LV outflow tract[22].

Overall 4D flow MRI resulted in a smoother flow field than echoPIV. It is likely that MRI captures a beat-averaged version of the instantaneous flow dynamics (as the flow data is reconstructed using multiple heart cycles and multiple averages), whereas echoPIV’s estimate captures the true time-resolved flow dynamics but is also partially corrupted by noise from the imaging and speckle tracking processes. Using physics-informed smoothing algorithms may help form a more accurate middle-ground between the two extremes in the future[23].

Another observation was that echoPIV captured finer flow details in the apex than 4D flow MRI. This is due partly to MRI’s low sensitivity to slow flow because of the chosen VENC setting in this study but also because of the higher spatial resolution of ultrasound close to the transducer (and higher temporal resolution in general). We also noted the flip side of the coin: in the basal region the ultrasound spatial resolution is lower (due to the sector-scan) resulting in seemingly wider filling jets than those captured with MRI.

EchoPIV’s improved temporal resolution also allowed for the assessment of dynamic changes in flow pattern over multiple cardiac cycles. In Figure 6 we observed that intra-beat variability in diastolic filling timing caused 4D flow MRI to largely underestimate the KE associated with early and late filling, as compared with echoPIV.

### Quantitative Comparison

The results of the quantitative comparative analysis of the velocity profiles and the derived flow parameters (Figure 4 & 5) show a good correspondence between echoPIV and 4D flow MRI. The Bland-Altman analysis showed MRI recorded higher velocities in the outflow tract during systole, while echocardiography measured higher velocities in the mitral inflow during diastole. It has previously been shown that echoPIV underestimated the filling and ejection velocities compared to pulsed-wave Doppler by 0.12m/s and 0.22m/s, respectively[6]. This indicates that both echoPIV and 4D flow MRI are similarly biased towards lower velocities than pulsed-wave Doppler. In addition, research comparing 4D flow MRI with traditional transthoracic pulse-wave Doppler echocardiography for the measurement of mitral inflow peak diastolic velocities reported that MRI underestimated peak diastolic velocities by 0.05 m/s (95% CI -0.37 to 0.46 m/s)[24]. Thus, the differences between echoPIV and 4D-flow MRI found in this study are comparable with previous inter-modality studies.

When compared to the velocity profiles, the derived regional energy parameters, KE and E·L exhibited a higher level of cosine similarity, which indicates good correspondence of temporal patterns. However, a higher NMAE was seen, indicating offsets in the absolute values, especially for E·L (calculated using derivatives of the velocity field) and more susceptible to noise and smoothing(Figure 3.F).

From the comparison of results from clinically feasible group and well-aligned group, we observed an increase in variance, indicating plane alignment is important for consistent estimation of the flow field. Therefore, sonographers should be critical of selecting the standard planes, especially for follow-up studies.

### Interpretation of Flow Features

To highlight the potential clinical benefit of HFR echoPIV we present two specific cases in Figure 7. Figure 7.A (Movie S.4)depicts the flow in a severely dilated left ventricle and reduced ejection fraction (LV ejection fraction 21%), characterized by a single large vortex that swirls blood inside the whole ventricle. This pathological condition is associated with a reduced relative energy loss (E·L/KE), as the KE is well preserved within the large vortex (Figure 7.B)[25].

**Figure 7.**
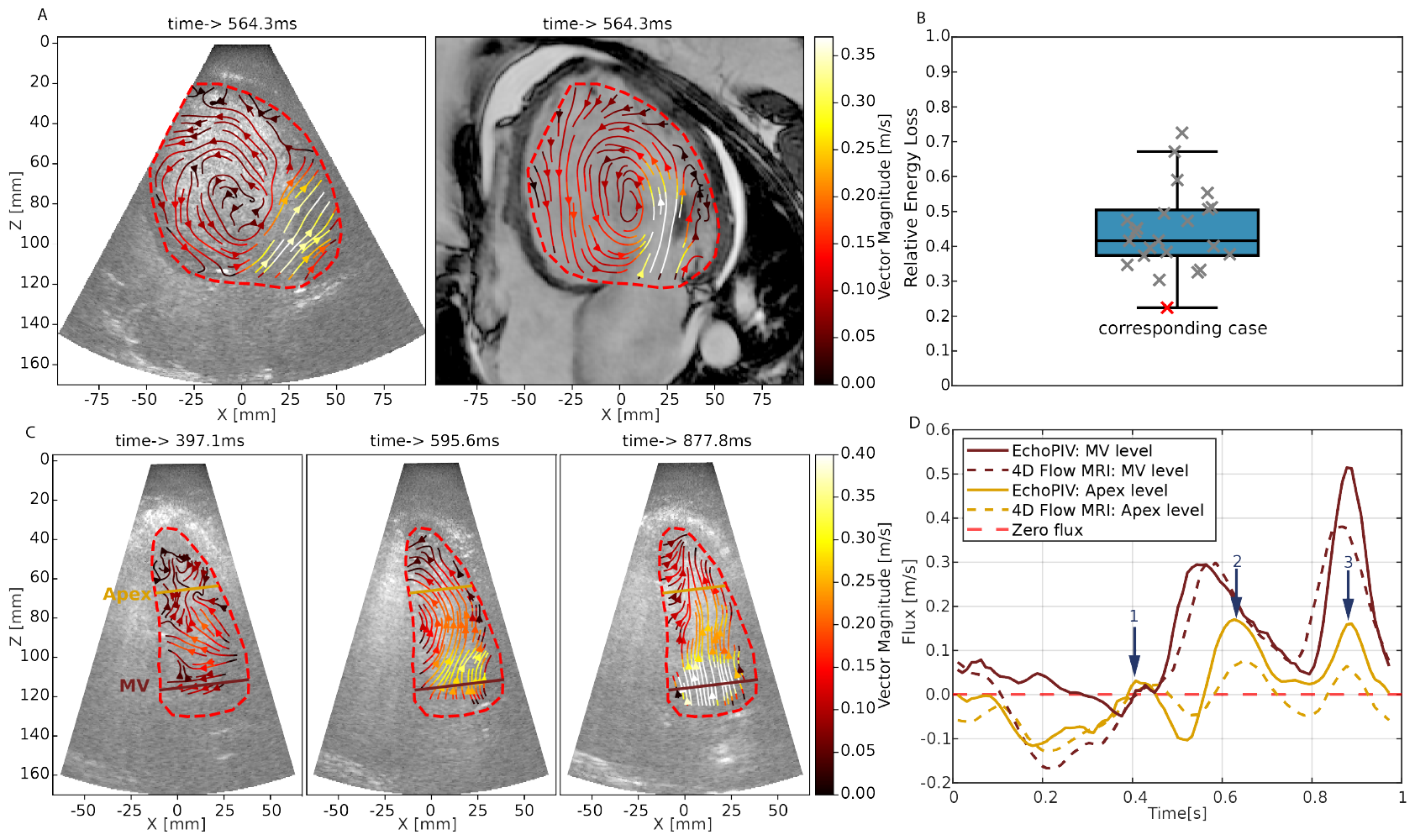
(A) Example of the flow pattern at peak early filling for a patient with severely dilated left ventricle and reduced ejection fraction. (B) Relative energy loss (energy Loss normalized by kinetic energy) for each included patient (the patient case shown in (A) is marked in red). (C) Example of the flow pattern at three filling phases for a patient with left bundle branch block. (D) Flux at base line and apical line (marked in (C)) for this patient.

Figure 7.C (Movie S.5) and D depict abnormal apical flow in a patient with left bundle branch block. The presence of an aberrant flow pattern is associated with irregular relaxation of the septal wall. Notably, this anomaly is not readily discernible through conventional echocardiographic measurements but is evident when examining the flow pattern.

### Challenges

To measure high-velocity flows within the heart, HFR ultrasound sacrifices some image quality compared to conventional focused ultrasound imaging. Furthermore, patients presenting with cardiac pathologies frequently exhibit comorbidities such as obesity or lung disease, which bring further challenges to acquiring sufficient-quality echocardiographic images.

The quality and accuracy of echocardiographic images are highly operator-dependent, potentially leading to variability in each acquisition. In this study, we used a probe with a relatively large footprint, which was difficult to fit between the ribs. This also made precise acquisition of the standard apical views difficult, especially for apical two-chamber view, where there are few landmarks to locate the plane. An example (Figure S.3) is shown in supplementary data, in which echocardiographic imaging plane is severely foreshortened and deviated from the standard apical 2 chamber view.

### Limitations

The focused patient cohort in this study might reduce the generalizability of our findings to a broader array of patient populations. Our research was confined to analyzing left ventricular flow, suggesting a potential avenue for future studies to apply echoPIV to other cardiac chambers. Furthermore, the comparison of the 2D velocity field from echoPIV with the projected 3D velocity field from 4D flow MRI might lead to discrepancies in flow measurement and interpretation.

### Conclusions

EchoPIV was feasible for 78% of the acquisitions. EchoPIV demonstrated the capability to accurately quantify flow within the LV, providing reliable spatial-temporal velocity distribution and magnitude measurement compared to 4D flow MRI as the reference standard. Both modalities have their benefits. MRI is more robust to patient-related factors that may influence the echocardiographic image quality. On the other hand, echoPIV, offers the advantage of capturing inter-beat variability and has a much higher temporal resolution.

## Data Availability

Data is available on reasonable request

## Acknowledgments

The work described in this article was funded by the Medical Delta program “Ultrafast Ultrasound for the Heart and Brain”; the project X-Flow of the research program Ultra-X-Treme (P17-32), which is financed by the Dutch Research Council (NWO); and the Dutch Heart Foundation (Hartstichting) as part of project number 03-004-2022-0044. We thank Dr. Ing Han Gho for his assistance in the contrast-enhanced ultrasound image acquisition.

## Supplementary Data

### High Frame Rate Echocardiography

HFR data were obtained with the setup described by Voorneveld et al.[16]. In brief, four repeated diverging-wave acquisitions with two alternating polarity transmits (pulse inversion) at two different angles (−7°, +7°) were used.

The saved RF data after pulse inversion summation was filtered at the second harmonic with a fourth-order Butterworth bandpass filter (2.6–3.8 MHz) to remove any residual fundamental frequency component and then beamformed onto a polar coordinate grid using the Verasonics software beamformer off-line. Singular Value Decomposition with automatic low-rank selection[19] and truncation was applied as a wall-filter.

### EchoPIV processing

The EchoPIV processing is described in detail in Voorneveld et al.[15][16]. In short, to estimate the displacements between subsequent frames, the image area was divided into equally sized blocks with overlap. Normalized cross correlation (NXCC) was then computed in the frequency domain for each block, and the peak of the correlation function, obtained through subpixel fitting, provided the displacement between the frames per block. The algorithm iteratively performed the blockwise NXCC step multiple times, using the displacements from the previous iteration to deform the target frame and reduce the displacement between frames towards zero. The window size was progressively reduced between iterations to enhance resolution and minimize displacement estimation bias.

Instead of coherent compounding of angled plane wave acquisitions, correlation compounding was employed. This involved performing blockwise normalized cross correlation between frames with similar angles and averaging the resulting correlation maps across different angles. Additionally, correlation averaging was applied across an ensemble of ten frames to further reduce noise.

Postprocessing steps included the application of a 2-D Gaussian spatial smoothing filter (with standard deviation of 0.5 and filter size of 3) and ensemble temporal moving average filter with a length of 10 frames to the computed velocity fields. Finally, the velocity data were scan converted to cartesian coordinates for visualization using the vector projectile imaging technique[26].

### Equations for agreement assessment and flow analysis

The Dice coefficient is computed as:

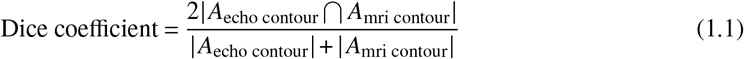

where *A* = Area enclosed by contour.

Cosine similarity employs the cosine of the angle between two profile vectors, yielding a similarity that ranges from -1 to 1. In our analysis, a cosine similarity close to 1 indicates a high degree of similarity between the temporal profiles obtained from the two imaging techniques.

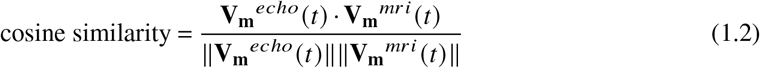

where ∥**V**_**m**_∥ is the Euclidean norm of vector **V**_**m**_(*t*), defined as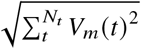. Note that Cosine Similarity represents the similarity in pattern shape, and can be high even if the norms are very different.

NMAE was used to quantify the average discrepancy between the velocity magnitudes measured by the two methods, providing insights into the accuracy of the absolute velocity measurements. It was calculated as the mean absolute difference between the measured (echoPIV) and the reference (MRI) velocities, normalized by their maximum.

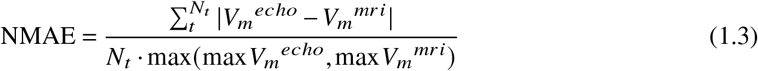

where *N*_*t*_ is the number of frames.

Kinetic energy was computed from the velocity vectors, 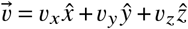, as

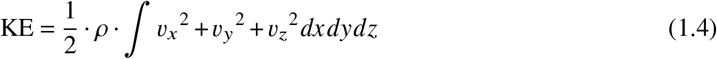

where ρ = 1060*kg*/*m*^3^ is the blood density, and the integral is over the region of interest. For a 2D acquisition, we assume an infinitely thin layer in through-plane direction (y-direction) and *v*_*y*_ = 0, thus Eq. 1.4 can be written as:

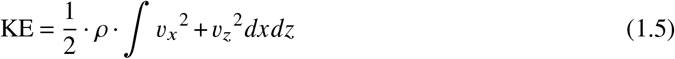

Rate of energy loss was estimated from the viscous dissipation, which considers the velocity gradients in the flow:

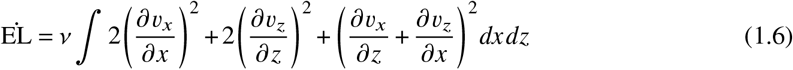

where *v* = 0.004*Pa*.*s* is the kinematic viscosity of blood. Circulation Γ within a vortex of blood flow was calculated as:

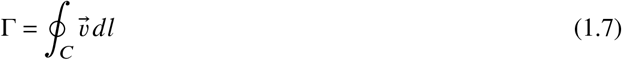

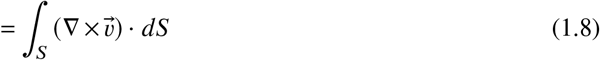

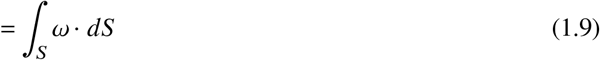

with the line integral taken over a closed circuit *C* around a vortex region in counter-clockwise sense.

### Extra figures

**Figure S.1.**
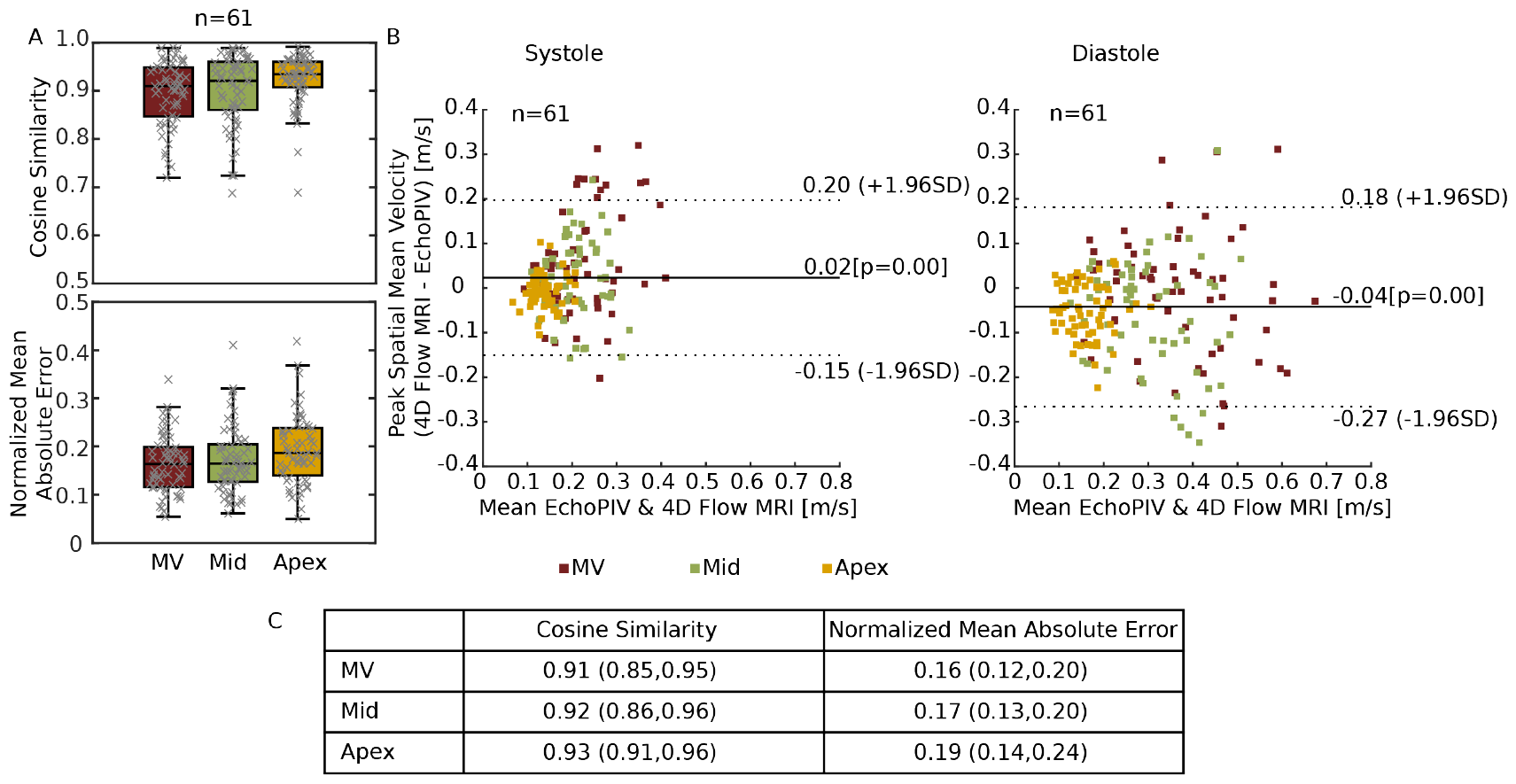
(A) Box plot of cosine similarity and normalized mean absolute error of the spatial-temporal velocity magnitude profile through lines at base, mid and apex level (for all feasible acquisitions). Black lines in the middle indicate median, boxes extend to 25th and 75th percentiles, and whiskers indicate the range excluding samples outside of 1.5 times the interquartile range. (B) Bland-Altman plot of the peak spatial mean velocity through lines at base, mid and apex level (as demonstrated in Fig 1.A) during systole and diastole. (C) Table summarizing the box plot in Fig. S.1.A, data are expressed in median(25th and 75th percentiles). MV=mitral valve

**Figure S.2.**
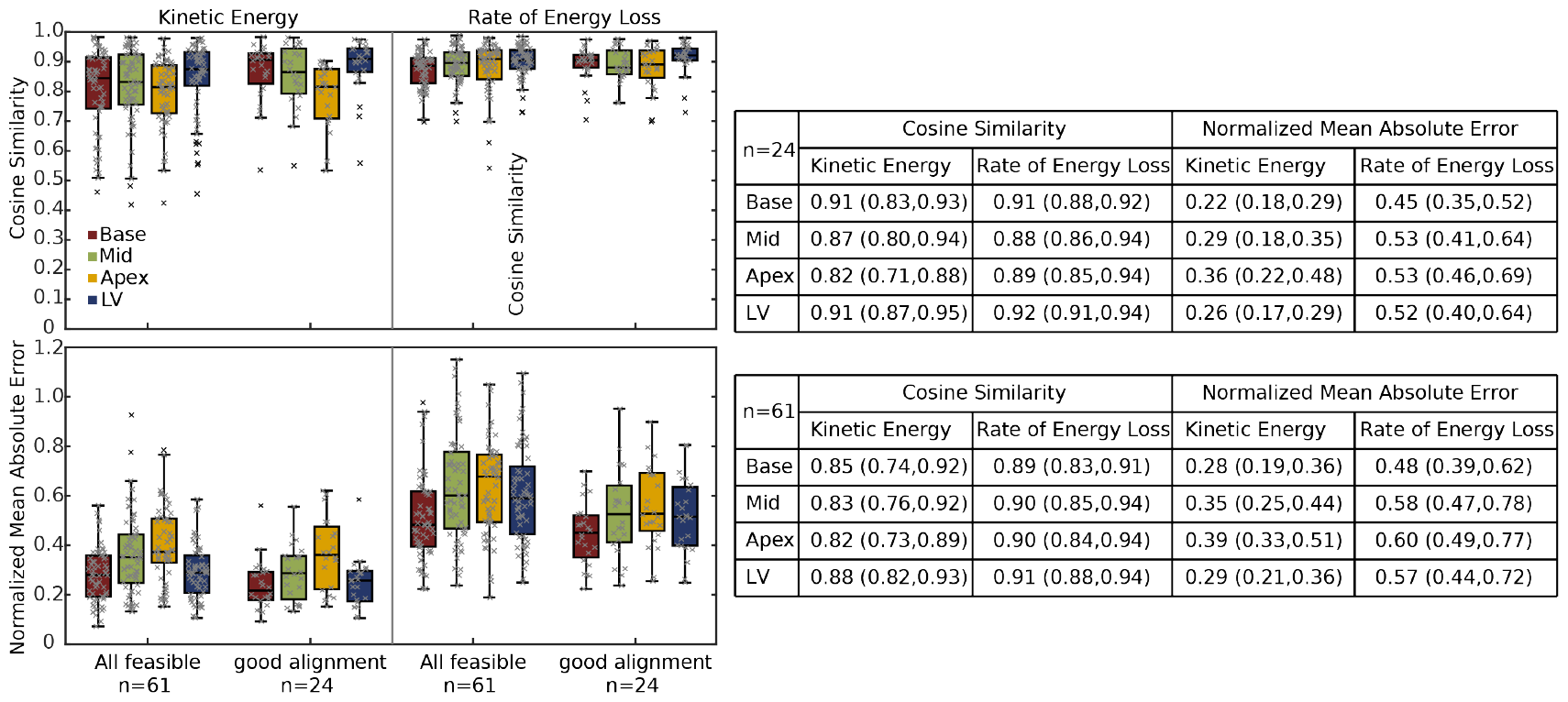
Box plot of cosine similarity and normalized mean absolute error of kinetic energy and energy loss rate in base, mid, apex regions and the whole left ventricle (LV) for all feasible acquisitions (n=61) and for the ones with good alignment (n=24). Black lines in the middle indicate median, boxes extend to 25th and 75th percentiles, and whiskers indicate the range excluding samples outside of 1.5 times the interquartile range. Data are expressed in median(25th and 75th percentiles)

**Figure S.3.**
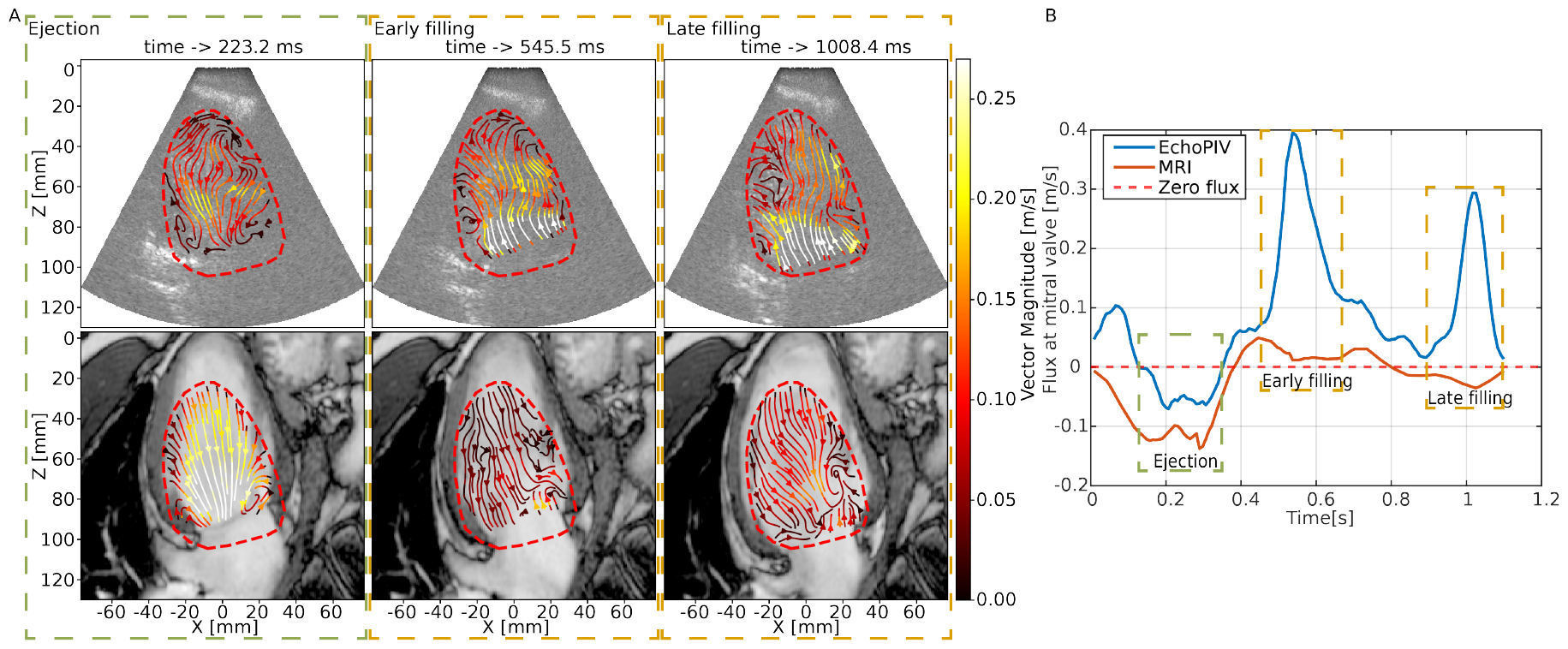
(A) Flow streamlines of EchoPIV (up) and MRI (bottom) at *t* = 223*ms* (ejection phase), *t* = 546*ms* (early filling) and *t* = 1008*ms* (late filling). (B) Flux at a plane at the level of mitral valve, with zero flux marking with red dashed line.

### Extra movies

Movie S.1. Pathlines of the flow field (EchoPIV on the left and MRI on the right) for the exemplary case shown in Figure 3.A.

Movie S.2. Vortex formation and propagation (EchoPIV on the left and MRI on the right) for the exemplary case shown in Figure 3.C.

Movie S.3. Pathlines of the flow field of the three distinct cardiac cycles for the example of inter-beat variability captured by echoPIV shown in Figure 6

Movie S.4. Pathlines of the flow field (EchoPIV on the left and MRI on the right) for the example of severely dilated left ventricle shown in Figure 7.A

Movie S.5. Pathlines of the flow pattern for the example of a patient with LBBB and have three apical level filling phases shown in Figure 7.C.

Movie S.6. Pathlines of the flow pattern for an example whose echo imaging plane was severely foreshortened and was thus excluded from flow comparison.

Movie S.7. Pathlines of the flow pattern for an example whose echo imaging plane was severely rotated from MRI imaging plane and was thus excluded from flow comparison.

Movie S.8. Pathlines of the flow pattern for an example whose echo imaging plane was severely foreshortened and rotated from MRI imaging plane and was thus excluded from flow comparison.

